# Prolonged Hospitalization Among Children Aged <5 years Admitted With Acute Gastroenteritis at Siaya County Referral Hospital, in Rural Western Kenya: 2010-2020

**DOI:** 10.1101/2025.06.06.25329108

**Authors:** Alex O. Awuor, Billy Ogwel, Bryan O. Nyawanda, Evans Apondi, Raphael Anyango, Sammy Khagayi, John Benjamin Ochieng, Erick Muok, Stephen Munga, George Ayodo, Victor Akelo, Sergon Kibet, Jason M. Mwenda, Umesh Parashar, Jacqueline E. Tate, Richard Omore

**Affiliations:** Kenya Medical Research Institute, Center for Global Health Research (KEMRI-CGHR), Kisumu, Kenya; Jaramogi Oginga Odinga University of Science and Technology, Bondo, Kenya; Liverpool School of Tropical Medicine, Kisumu, Kenya; WHO Country Office for Kenya, Nairobi, Kenya; WHO Regional Office for Africa (WHO/AFRO), Brazzaville, Republic of the Congo; Division of Viral Diseases, US Centers for Disease Control and Prevention, Atlanta, GA, USA

**Keywords:** Gastroenteritis, prolonged hospitalization, children

## Abstract

**Background:** Acute gastroenteritis (AGE) causes substantial morbidity and mortality in children <5 years old accounting for 9 million hospitalizations. Prolonged hospitalization can cause dire consequences to the patient and healthcare system. However, data on factors associated with prolonged hospitalization for AGE in developing countries are limited.

**Objectives:** We aim to describe trends and assess factors associated with prolonged hospitalization among children <5 years admitted with AGE in western Kenya.

**Methods:** Children with AGE (≥3 loose stools and/or ≥1 episode of unexplained vomiting with loose stool within 24 hours) hospitalized at Siaya County Referral Hospital from January 2010 through December 2020 were included. Prolonged hospitalization was defined as admission for ≥5 days. Trends of prolonged AGE hospitalizations were assessed using Cochran-Armitage trend test while factors associated with prolonged hospitalization for AGE were determined by unconditional logistic regression.

**Results:** Of the 12,546 all-cause admissions among children <5 years, 2,271(18.1%) children had AGE; 681 (32.8%) had prolonged hospitalization. There was a significant difference in the prevalence of prolonged hospitalization over time, with a peak in 2010 (42.8%] and a low in 2016 (10.8%). Older children (12-23 months: (adjusted Odds ratio [aOR]: 0.69; 95% confidence interval [95% CI]: 0.49-0.97)) and those who vomited everything (aOR: 0.69; 95% CI: 0.52-0.90) were less likely to have prolonged hospitalization. Children who had a bulging fontanelle (aOR: 3.21; 95% CI: 1.12-9.20) or chest in drawing (aOR: 1.49; 95% CI: 1.02-2.18) or were severely stunted (aOR: 2.67; 95% CI: 1.89-3.79) or severely wasted (aOR: 2.34; 95% CI: 1.65-3.30) were more likely to have prolonged hospitalization.

**Conclusion:** Children with severe diarrheal illness with malnutrition are at high risk of prolonged hospitalization. Targeted interventions such as increased clinical and diagnostics monitoring for at-risk children with AGE may need to be prioritized to reduce possible prolonged hospitalization.

## Introduction

Pediatric hospitalizations for acute gastroenteritis (AGE) impose a substantial disease-related burden, with financial and non-monetary repercussions affecting both patients and health systems [1,2]. This impact extends to the broader family, encompassing direct and indirect costs, as well as influencing social, educational, and quality of life indicators (1). The evaluation of the burden of hospitalization relies on assessing the length of hospital stays as a key metric. This information serves as a crucial factor in guiding planning initiatives, allocating resources effectively, and fostering continuous improvements in the quality of care provided (2). Prolonged hospitalization has adverse outcomes for both the patients and the hospital, including high complications, increased risk of hospital-acquired infections, high costs of treatment, and decreased patient satisfaction (3–5).

While several studies have explored the duration of hospitalization for various illnesses (6–8), there is limited research on prolonged hospitalization due to AGE. AGE is generally defined as diarrhea or vomiting (or both), it may be accompanied by fever, abdominal pain, and anorexia with diarrhea duration <7 days.(9) AGE causes an estimated 1.7 billion episodes annually among children aged <5 years globally, leading to 124 million clinic visits, 9 million hospitalizations, and 1.34 million deaths, with more than 98% of these deaths occurring in the developing world (10,11). Moreover, despite the introduction of vaccination against rotavirus, the leading etiology of AGE, up to 10% of all hospitalizations of children <5 years of age are still due to AGE (12). Specifically in Kenya, approximately 21% of an estimated 1.5 million AGE episodes are hospitalized annually (13). This AGE-associated burden of hospitalization in children accompanied by a dearth of studies in developing nations on duration of hospitalization necessitates the need to understand factors driving prolonged hospitalization (10,11), which could help to identify those at risk to inform targeted interventions and design appropriate prevention strategies that can improve both patient outcomes and enhance healthcare system efficiency. Thus, we used data from rotavirus inpatient surveillance to establish trends and factors associated with prolonged hospitalization for AGE.

## Methods

### Study setting and population

The study area is Siaya County in rural western Kenya within the catchment area of Siaya County Referral Hospital (SCRH), a pivotal regional healthcare facility catering to a predominantly rural and semi-rural population. Notably, the hospital boasts a pediatric bed capacity of 60. This current analysis is nested in a broader framework of rotavirus surveillance, constituting a prospective study conducted within the hospital.

The study setting and population have been previously described in detail elsewhere(13,14). This community grapples with a heightened under-five mortality ratio, primarily attributed to infectious diseases(15). The high prevalence of endemic malaria [16], coupled with challenges posed by AGE and an equally high prevalence of HIV, underscores the complex health landscape of the region (17).

### Study Design and Procedures

Prospective hospital-based rotavirus surveillance enrolled children aged 0–59 months who were residents of Karemo Health and Demographic Surveillance System (HDSS) and hospitalized at the in-patient department of SCRH with AGE; defined as ≥3 looser than normal stools and/or ≥ 1 episode of unexplained vomiting followed by loose stool within a 24-h period beginning within the 7 days before seeking healthcare (13). We defined prolonged hospitalization as an admission duration of ≥5 days in the ward. We used the 75^th^ percentile cut-off point of the AGE hospitalization duration to determine prolonged hospitalization consistent with previous studies looking at length of stay for other medical conditions (7,18–20).

Trained health facility recorders approached all eligible paediatric patients, explained the study, and administered a questionnaire on demographics to their caretakers after obtaining informed consent. A study clinician then examined these patients and administered the standardized questionnaire to their parent/caretaker to gather information about symptoms, medical history, laboratory investigations, diagnosis, treatment and outcome of hospitalization. During data collection, built-in software in the electronic questionnaire with built-in checks and controls ensured quality control.

Our study focussed on children aged <60 months hospitalized with AGE at SCRH from January 1, 2010 to December 31, 2020 when the rotavirus surveillance stopped. We included all children admitted with AGE during this period except those who died while hospitalized and that missing discharge information.

### Statistical analysis

Descriptive statistics were presented in percentages and counts using frequency tables. Proportion of admissions due to AGE was calculated by dividing the number of AGE cases by the number of all-cause admissions at SCRH who were residents of Karemo HDSS during the study period. We used Cochran-Armitage trend test to assess the trends in proportion of prolonged hospitalization for AGE over time. We compared clinical, demographic, epidemiological, and medical management factors of AGE cases with prolonged hospitalization (≥5 days) versus those who had non-prolonged hospitalization (<5 days) using logistic regression. We assessed collinearity across variables using Cramer’s V statistic and if present, then only the variable with the strongest association with prolonged hospitalization was considered for the multivariable regression. Variables with P-value < 0.2 from bivariate analysis were included in the multivariable model. We reported the adjusted Odds Ratios (aOR) and the accompanying 95% confidence intervals (95%CI). For all analyses, a p-value less than 0.05 was considered statistically significant. Data analysis was done using Stata/SE 16.0

### Ethical Considerations

The rotavirus inpatient surveillance was reviewed and approved as part of the HDSS protocol by the Kenya Medical Research Institute Scientific and Ethical Review Unit (SSC # 1801) and Institutional Review Board of CDC (CDC IRB #3308). Caregivers provided written consent before study procedures.

## Results

### Participants’ characteristics

From January 1, 2010 to December 31, 2020, 12,546 all-cause admissions occurred among children <5 years in SCRH and of whom, 2,271 (18.1%) children <5 years were admitted with AGE. Of the 2,271 patients admitted with AGE, 195 (8.5%) were excluded from the analysis; 34 with missing dates of discharge and 161 deaths. Of the 2,076 AGE cases that were analyzed, slightly more than half were males (1158/2076 (55.8%)). The distribution of age among those who had AGE were as follows 0-11 months (1251 {60.3%}), 12-23 months (544, {26.2%}) and 24-59 months (281 {13.5%}). Among the admitted children, 681 (32.8%) had prolonged AGE hospitalization while 1365 (67.2%) did not (Fig 1).

**Figure 1:**
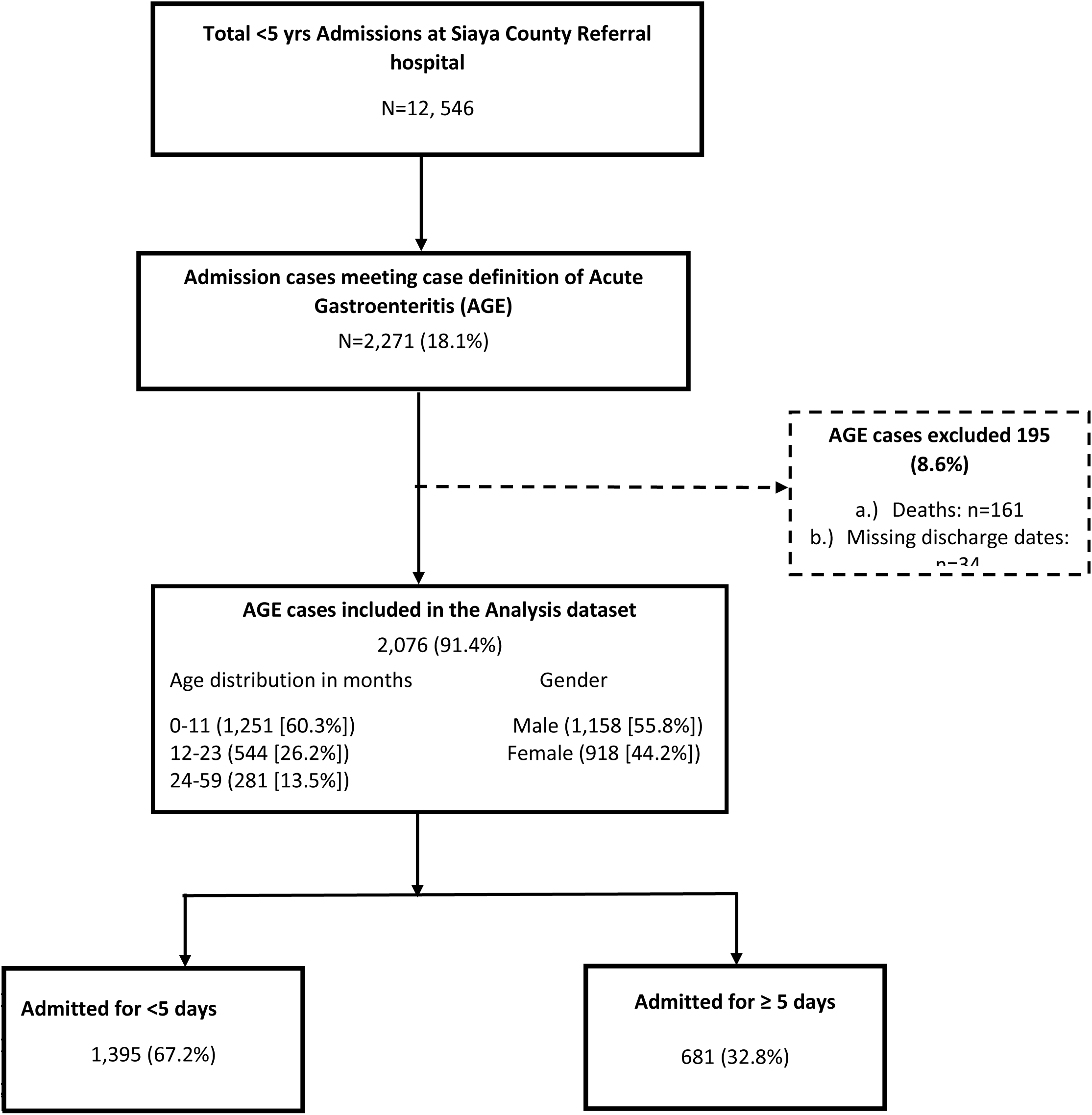
Flow diagram of hospitalization duration among children aged <5 years admitted with gastroenteritis at Siaya County Referral Hospital, western Kenya 2010–2020

### Trends of prolonged hospitalization among children aged <5 years admitted with AGE

We observed a notable decline in the number of hospitalizations attributable to AGE over the years, with the highest count recorded in 2010 (608 cases) and the lowest in 2017 (34 cases). However, a subsequent resurgence of hospitalized AGE cases occurred in 2018, peaking in 2020 with 86 cases. The prevalence of prolonged hospitalization for AGE exhibited significant variation throughout the years (p<0.0001), reaching its highest in 2010 at 42.8% and its lowest in 2016 at 10.8% (Fig 2).

**Figure 2:**
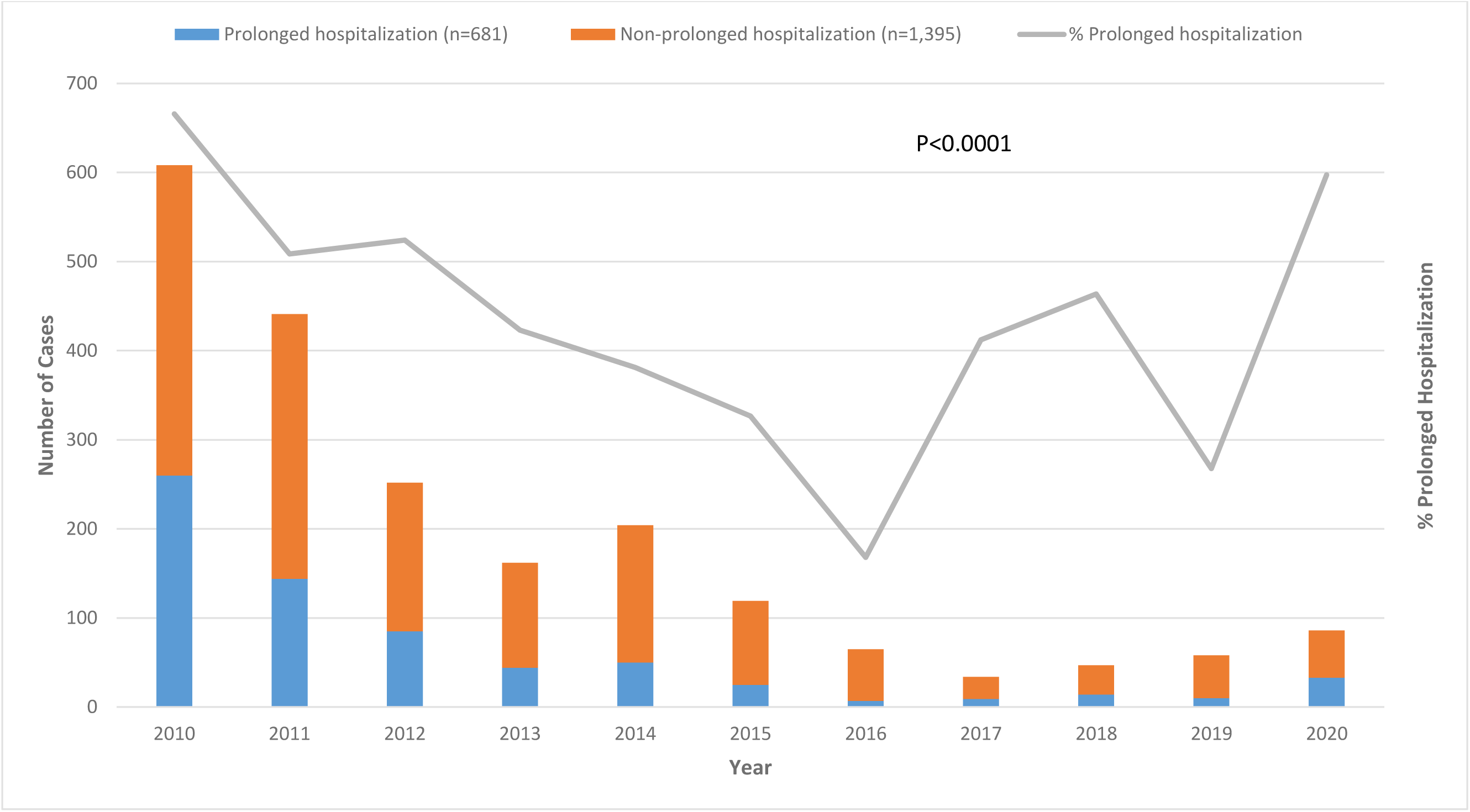
Trends of hospitalization across the years among children <5 years admitted with AGE, 2010-2020

The median duration of hospitalization for AGE was 3 days, with an interquartile range (IQR) of 2-5 days. The characteristics of AGE cases stratified by duration of hospitalization are shown Table 1. Children who had prolonged hospitalization for AGE were younger than those who did not (median age in months [IQR]: 9.5 [5.7-16.3] vs 10.1 [6.3-17.6], p= 0.0094). Moreover, patients with prolonged hospital stay had a higher respiratory rate (Median Respiratory Rate [IQR] 43 [36-56] vs 40 [34-50], p= <0.0001) and more severe disease (median Vesikari Score [IQR]: 11 [8-12] vs 10 [8-12], p=<0.0001) compared to those who did not. Additionally, children who vomited everything, had convulsions, were lethargic, had weight loss, who responded to voice command or mild pain, had sunken eyes, had decreased skin turgor, slow capillary refill (>2 seconds), had sunken fontanelle, had intravenous fluids or ORS administered, were dehydrated, had stunting or wasting, were underweight, had a prior admission, had stridor, had chest in drawing or had nasal flaring were significantly more likely to have prolonged hospitalization than children with shorter hospitalizations (p<0.05).

**Table 1:** Factors associated with prolonged hospitalization among children < 5years admitted with AGE at SCRH: 2010-2020.

| Characteristics | Hospitalization |  |  | Bivariate |  | Multivariable* |
| --- | --- | --- | --- | --- | --- | --- |
|  | Prolonged: ≥ 5 days<br>(n=681) | Non-prolonged: < 5 days<br>(n=1,395) | P-Value | unadjusted OR(95% CI) | p-value | Adjusted OR(95% CI) |
| Median age months[IQR] | 9.5 [5.7-16.3] | 10.1 [6.3-17.6] | **0.0094** | 0.99 [0.98-0.99] | **0.05** |  |
| Age Categories |  |  |  |  |  |  |
| 0-11 months | 434 (63.7) | 817 (58.6) | 0.075 | Ref |  | Ref |
| 12-23 months | 161 (23.7) | 383 (27.5) |  | 0.79 [0.64-0.98] | **0.035** | **0.69 [0.49-0.97]** |
| 24-59 months | 86 (12.6) | 195 (13.9) |  | 0.83 [0.63-1.10] | 0.191 | 0.99 [0.62-1.59] |
| Gender: Male | 374 (54.9) | 784 (56.2) | 0.581 | 0.95 [0.79-1.14] | 0.581 |  |
| **Vital signs** |  |  |  |  |  |  |
| Median Temperature[IQR] | 37.2 [36.6-38.0] | 37.1 [36.6-38.0] | 0.5512 | 1.02 [0.98-1.06] | 0.269 |  |
| Median Oxygen Saturation[IQR] | 97 [93-98] | 97 [95-98] | **0.0014** | 0.97 [0.96-0.99] | **<0.001** |  |
| Median Respiratory Rate [IQR] | 43 [36-56] | 40 [34-50] | **<0.0001** | 1.02 [1.01-1.03] | **<0.001** | **1.02 [1.01-1.03]** |
| Median Pulse [IQR] | 146 [132-163] | 145 [130-162] | 0.3654 | 1.00 [0.99-1.01] | 0.356 |  |
| **Symptoms** |  |  |  |  |  |  |
| Vomiting | 514 (75.5) | 1,065 (76.3) | 0.664 | 0.95 [0.77-1.18] | 0.664 |  |
| Vomits Everything | 244 (47.9) | 578 (55.1) | **0.008** | 0.75 [0.61-0.93] | **0.008** | **0.69 [0.52-0.90]** |
| Convulsions | 108 (16.3) | 272 (20.3) | **0.032** | 0.76 [0.60-0.98] | **0.032** |  |
| Lethargic | 349 (55.0) | 584 (48.7) | **0.011** | 1.29 [1.06-1.56] | **0.011** |  |
| Unconscious | 74 (11.2) | 132 (9.9) | 0.363 | 1.15 [0.85-1.55] | 0.363 |  |
| Fever | 580 (86.8) | 1,178 (87.5) | 0.661 | 0.94 [0.71-1.23] | 0.661 |  |
| Malaria | 206 (32.2) | 462 (36.1) | 0.09 | 0.84 [0.69-1.03] | 0.09 |  |
| Lost weight | 442 (69.5) | 708 (59.6) | **<0.0001** | 1.54 [1.26-1.90] | **<0.001** |  |
| Unable to drink | 32 (6.4) | 43 (4.8) | 0.212 | 1.35 [0.84-2.16] | 0.213 |  |
| **AVPU** |  |  |  |  |  |  |
| Alert | 646 (95.3) | 1,356 (98.0) | **0.002** | Ref |  |  |
| Voice | 18 (2.6) | 19 (1.4) |  | 1.99 [1.04-3.81] | **0.039** |  |
| Pain | 12 (1.8) | 7 (0.5) |  | 3.60 [1.41-9.18] | **0.007** |  |
| Unresponsive | 2 (0.3) | 1 (0.1) |  | 4.20 [0.38-46.38] | 0.242 |  |
| Restless | 158 (23.4) | 313 (22.7) | 0.732 | 1.04 [0.84-1.29] | 0.732 |  |
| Convulsing Now | 14 (2.1) | 15 (1.1) | 0.073 | 1.94 [0.93-4.04] | 0.078 |  |
| Sunken Eyes | 210 (31.0) | 352 (25.4) | **0.007** | 1.32 [1.08-1.62] | **0.007** |  |
| Skin Pinch |  |  |  |  |  |  |
| Normal | 414 (61.3) | 1,021 (73.4) | **<0.0001** | Ref |  |  |
| Slowly | 208 (30.8) | 322 (23.2) |  | 1.59 [1.29-1.96] | **<0.001** |  |
| Very Slowly | 53 (7.9) | 47 (3.4) |  | 2.78 [1.85-4.19] | **<0.001** |  |
| Capillary Refill (>2 seconds) | 123 (18.5) | 186 (13.9) | **0.007** | 1.41 [1.10-1.81] | **0.007** |  |
| Bulging Fontanelle | 12 (2.3) | 11 (1.2) | 0.1 | 1.97 [0.86-4.50] | 0.107 | **3.21 [1.12-9.20]** |
| Sunken Fontanelle | 127 (21.2) | 199 (16.7) | **0.019** | 1.34 [1.05-1.72] | **0.02** |  |
| Red Eyes | 2 (0.4) | 3 (0.3) | 0.843 | 1.20 [0.20-7.20] | 0.843 |  |
| IVF given | 238 (35.2) | 566 (41.0) | **0.011** | 0.78 [0.65-0.95] | **0.011** |  |
| ORS given | 75 (22.7) | 278 (32.6) | **0.001** | 0.61 [0.45-0.81] | **0.001** |  |
| Dehydration Status |  |  |  |  |  |  |
| No dehydration | 393 (57.7) | 925 (66.3) |  | Ref |  |  |
| Some dehydration | 230 (33.8) | 412 (29.5) |  | 1.31 [1.08-1.60] | 0.007 |  |
| Severe dehydration | 58 (8.5) | 58 (4.2) | **<0.0001** | 2.35 [1.61-3.45] | **<0.001** |  |
| Vesikari Score |  |  |  |  |  |  |
| Mild | 90 (13.2) | 199 (14.3) | **0.029** | Ref |  |  |
| Moderate | 245 (36.0) | 573 (41.1) |  | 0.95 [0.71-1.26] | 0.705 |  |
| Severe | 346 (50.8) | 623 (44.7) |  | 1.23 [0.93-1.63] | 0.153 |  |
| Stunting |  |  |  |  |  |  |
| Normal | 371 (55.1) | 968 (69.8) | **<0.0001** | Ref |  | **Ref** |
| Moderate | 117 (17.4) | 223 (16.1) |  | 1.37 [1.06-1.76] | **0.015** | 1.41 [0.99-2.01] |
| Severe | 185 (27.5) | 195 (14.1) |  | 2.48 [1.96-3.12] | **<0.001** | **2.67 [1.89-3.79]** |
| Wasting |  |  |  |  |  |  |
| Normal | 405 (61.4) | 1,017 (74.5) | **<0.0001** | Ref |  | **Ref** |
| Moderate | 89 (13.5) | 166 (12.2) |  | 1.35 [1.02-1.79] | 0.039 | 1.39 [0.93-2.08] |
| Severe | 166 (25.1) | 181 (13.3) |  | 2.30 [1.81-2.93] | **<0.001** | **2.34 [1.65-3.30]** |
| Underweight |  |  |  |  |  |  |
| Normal | 344 (51.8) | 952 (69.4) | **<0.0001** | Ref |  |  |
| Moderate | 103 (15.5) | 234 (17.1) |  | 1.22 [0.94-1.58] | 0.141 |  |
| Severe | 217 (32.7) | 186 (13.5) |  | 3.22 [2.56-4.07] | **<0.001** |  |
| Prior Admission | 15 (3.3) | 14 (1.6) | **0.035** | 2.17 [1.04-4.55] | **0.039** |  |
| Sought Care | 382 (61.8) | 708 (60.3) | **0.521** | 1.07 [0.87-1.30] | 0.521 |  |
| Chest indrawing | 145 (21.8) | 174 (13.0) | **<0.0001** | 1.87 [1.47-2.39] | **<0.001** | **1.49 [1.02-2.18]** |
| Stridor | 39 (6.1) | 43 (3.6) | **0.011** | 1.77 [1.13-2.75] | **0.012** |  |
| Nasal flaring | 149 (23.4) | 191 (15.8) | **<0.0001** | 1.62 [1.28-2.06] | **<0.001** |  |
| Median Diarrhea days [IQR] | 3 [2-5] | 3 [2-4] | **0.0002** | 1.09 [1.04-1.15] | **0.001** | **1.07 [1.00-1.15]** |
| Median Diarrhea episodes [IQR] | 4 [3-5] | 4 [3-5] | 0.806 | 0.99 [0.99-1.00] | 0.508 |  |
| Median Vomit episodes [IQR] | 3 [1-4] | 3 [1-4] | 0.0982 | 0.99 [0.96-1.02] | 0.546 |  |
| Median Vesikari Score [IQR] | 11 [8-12] | 10 [8-12] | **<0.0001** | 1.04 [1.01-1.07] | **0.011** |  |

\* Variables that had a P-value <0.2 were included in the multivariable model: Age, Oxygen Saturation, Respiratory Rate, Vomits Everything, Vomits Everything, Convulsions, Lethargic, Malaria, Lost weight AVPU scale, Convulsing Now, Sunken Eyes, Skin Pinch, Capillary Refill, Bulging Fontanelle, Sunken Fontanelle, Red Eyes, IVF given, ORS given, Stunting, Wasting, Prior Admission, Chest indrawing, Stridor, Nasal flaring and Diarrhea days
\*\* Underweight was dropped because it was collinear with wasting

### Factors associated with prolonged hospitalization for AGE by multivariable analysis

From our multivariable analysis, we observed that a unit increase in respiratory rate increased the probability of prolonged hospitalization by 2% (adjusted odds ratio [aOR]=1.02, 95% confidence interval [95%CI] 1.01-1.03). Additionally, a unit increase in diarrhea days increased the probability of prolonged hospitalization by 7% (aOR=1.07, 95%CI 1.00-1.15). Children who had bulging fontanelle were 3 times more likely to have prolonged hospitalization (aOR=3.21, 95%CI 1.12-9.20) compared to those who did not. Furthermore, children who were severely stunted (aOR=2.67, 95%CI 1.89-3.79) and severely wasted (aOR=2.34, 95%CI 1.65-3.30) were more likely to have prolonged hospitalization compared to those who were normal. Children presenting with chest in drawing were more likely to have prolonged hospitalization (aOR=1.49, 95%CI 1.02-2.18) compared to those who did not. Contrarily, we observed age (12-23 months: aOR=0.69, 95%CI 0.49-0.97) and vomiting everything (aOR=0.69, 95%CI 0.52-0.90) to be negatively associated with prolonged hospitalization (Table 1).

The distribution of clinical symptoms and signs were similar among those with prolonged and non-prolonged hospitalization; however, signs of malnutrition were more common among those with prolonged hospitalization.

## Discussion

In our study of the trend and factors associated with prolonged hospitalization among pediatric patients with AGE in rural Western Kenya, we observed a relatively high prevalence of prolonged hospitalization with a significant variation throughout the years. Younger children aged <1 year were more likely to have prolonged hospitalization than older children and severity of diarrheal disease, malnutrition (stunting and wasting), and co-infection (signs of respiratory distress and bulging fontanel) were independently associated with prolonged hospital duration for AGE.

Even though we observed a decline in AGE hospitalizations over the study period, the overall prevalence of prolonged hospitalization was relatively high ∼32%. Analyzing the trend of a disease is crucial for evaluating the effectiveness of interventions and understanding fluctuations in its prevalence. Our examination revealed a high prevalence of prolonged hospitalization associated with AGE in 2010. Subsequently, there was a discernible decline leading up to 2016, followed by a subsequent increase until 2018. Another decline was observed in 2019, succeeded by an increase in 2020. While the proportion of AGE hospitalization started declining from 2010, the introduction of rotavirus vaccine in 2014 could have exacerbated this decline as the vaccine has been shown to reduce severity of gastroenteritis infection cases caused by the virus (21). The increasing prevalence of prolonged hospitalization towards the end of the study period may be partially attributable to rotavirus vaccine introduction. Rotavirus hospitalizations tend to be of short duration (<5 days) in rural western Kenya— as demonstrated by Omore et al. in which showed a median duration of 2days for rotavirus hospitalization(22). A decrease in shorter hospitalizations would result in a higher proportion of remaining AGE hospitalizations that are prolonged.

Studies have consistently shown a decrease in admissions related to rotavirus infections following the widespread implementation of the vaccine [15]. This suggests a positive correlation between the vaccine intervention and the observed decline in hospitalizations, emphasizing its potential impact on mitigating the impact of AGE. Additionally our study observed similar findings in the median length of hospitalization with studies conducted in Australia and Ethiopia which showed a median length of hospitalization of 3 days (20,23,24). However, this finding was lower than the findings from studies conducted in Saudi Arabia (median 6 days) (25), Tunisia (median 8 days) (26) and Uganda (median 5 days) (27). This variation of length of hospitalization is expected because of differences in the type of health facilities setups across the world. For instance, the efficiency of the health care system that could in turn improve quality of care. Furthermore, timing of the studies within the global COVID-19 pandemic could influence the length of hospitalization including in 2020 for our study (28).

The age of children emerged as a significant socio-demographic factor in our study, playing a pivotal role in influencing the recovery outcomes of patients with AGE. Specifically, infants exhibited the highest proportion of cases with prolonged hospitalization in comparison to their counterparts aged over 1 year. This finding is consistent to a study conducted by Weiss et al. in USA, which also showed that infants under 1 year of age constituted the largest proportion of total pediatric hospitalizations at 76.7% (29). This could be associated with lowered immunity in young infants as their body immunity to resist or fight infection is still immature and might lead to severe disease resulting in prolonged hospitalization. Understanding the relationship between age and duration of hospitalization can be instrumental in tailoring interventions and medical care to address the specific needs of different age groups affected by AGE.

Our study examined the nutritional status of children admitted to the hospital with AGE. We identified severe wasting and stunting as significant predictors of prolonged hospitalization in these children. The relationship between malnutrition and gastroenteritis forms a vicious cycle, wherein malnourished children may experience persistent diarrhea, ultimately leading to prolonged hospitalization [24, 25]. Khagayi et al conducted a study revealing that malnutrition appears to diminish rotavirus vaccine effectiveness in preventing severe cases of AGE. This underscores the interconnectedness of nutritional status and the efficacy of vaccines. To optimize the benefits of vaccines, particularly in the context of gastroenteritis, it is crucial to address and enhance both rotavirus uptake and nutritional well-being [26]. Moreover, in a study by Nasrin et al, it was observed that an episode of moderate to severe diarrhea during the first 5 years of life was associated with a higher likelihood of stunting within 2–3 months after the episode [27]. Another study conducted by Farzana et al. in Bangladesh showed that among children with diarrhea, 28%,15% and 15% were malnourished, stunted and wasted, respectively (30). Similarly, a study conducted by Negasa in Ethiopia observed that malnourished children are more susceptible to diarrhea and other infections. Repeated diarrhea infections contribute to long-term gut damage that prevents nutrient absorption and immune system function, even when children eat healthy foods (31).This emphasizes the long-term consequences of malnutrition and gastroenteritis, highlighting the importance of preventive measures and interventions to mitigate the impact on children’s growth and development. Efforts to improve nutritional status and prevent malnutrition can play a pivotal role in minimizing the severity and duration of gastroenteritis-related hospitalizations. These findings are also similar to a study conducted by Assefaw et al, which showed the probability of recovery in malnourished children with severe wasting was reduced by 36% compared to well-nourished children (32).

Our study revealed a noteworthy association between the severity of diarrheal disease and co-infections marked by signs of respiratory distress and a bulging fontanel with prolonged hospitalization. This finding emphasizes the significant influence that both the intensity of severe AGE and concurrent infections can exert on the duration of hospitalization. These findings are consistent with a study conducted in Ethiopia (32) which showed that chest in drawing was more likely associated with prolonged hospitalization. These symptoms and signs are clinical indicators of severe illness and probably leading to severe disease progression that might require prolonged hospitalization to receive treatment (32). It emphasizes the interconnected nature of these factors, show casing their collective effect on the overall severity and management of patients, thus contributing to a more comprehensive understanding of the clinical implications.

Conversely, our study revealed an intriguing finding: children diagnosed with AGE who exhibited a symptom of vomiting everything were more likely to experience non-prolonged hospitalization. This observation serves as a protective factor against the primary outcome of interest, namely prolonged hospitalization. The rationale behind this phenomenon can be elucidated by considering the association between vomiting and a viral etiology, particularly rotavirus-induced diarrhea. Viral infections, such as those caused by rotavirus, often manifest with severe symptoms, leading to rapid dehydration. Consequently, prompt medical intervention involving the administration of intravenous fluids accelerates the recovery process, resulting in a shorter hospital stay for these infants. In essence, the association between vomiting and non-prolonged hospitalization underscores the potential effectiveness of timely and targeted interventions in managing severe viral gastroenteritis, thus mitigating the risk of extended hospital stays among affected children.

Understanding these predictors of prolonged hospitalization is the significance of this study. These findings could help policy makers and clinicians attending to children admitted with AGE in developing targeted interventions such as increased and targeted clinical and diagnostics monitoring for children seeking care for AGE and presenting with these conditions may need to be prioritized to reduce possible prolonged hospitalization.

Our study had a limitation of qualitative data to elicit caretaker factors and system factors associated with prolonged hospitalization. Additionally, we did not examine etiologies of AGE and how it changed over time, which could further have explained some of the factors associated with prolonged hospitalization. Moreover, there could have been changes in clinical management over time that influenced length of hospitalization.

## Conclusions

Our study findings revealed that pediatric populations with severe diarrheal illness accompanied with malnutrition features are at increased risk of prolonged hospitalization. Clinicians managing children with AGE should be very attentive to clinical features associated with prolonged hospitalization to offer prompt and high quality care to these children at high risk of prolonged hospitalization. Early identification and appropriate management of the signs of severe gastroenteritis may have the greatest effect on health outcomes. Strengthening clinical practices and nursing care according to national guidelines should be a priority for gastroenteritis management to shorten the duration of hospital stay.

Targeted interventions such as increased clinical and diagnostics monitoring for children seeking care for AGE presenting with these conditions may need to be prioritized to reduce possible prolonged hospitalization are recommended and conducting prospective studies to include primary data from caregivers can help understand caretaker factors associated with prolonged hospitalization.

## Data Availability

Data used in this study belongs to Kenya Medical Research Institute and will be available upon request to the corresponding author and according to institutional guidelines.

## Acknowledgement

This study includes data generated from the Rotavirus surveillance at SCRH. Therefore, we appreciate the contributions and support from the Siaya County referral hospital where the study was conducted and Kenya Ministry of Health. We appreciate the contributions and support from study staff and leadership including WHO county office for Kenya, WHO Regional Office Africa and Division of Viral Diseases, US Centers for Disease Control and Prevention. We wish to thank the study participants and their families for their voluntary participation. The findings and conclusions in this paper are those of the authors and do not necessarily represent the official position of KEMRI, US CDC, or any of the collaborating institutions.

## Conflict of Interest

The authors declare no conflict of interest.

## Disclosure

The findings and conclusions in this paper are those of the authors and do not necessarily represent the official position of, KEMRI, the US Centres for Disease Control and Prevention, or other collaborators.

## Author Contributions

A.O.A and BO conceived the idea of the study. A.O.A took a key leading role by substantially contributing to the conception and design of the work, leading in writing the manuscript. B.O. played a key role in helping in the analysis and interpretation of data for the work. All authors critically reviewed the manuscript and approved the final version of the manuscript for submission.

## Funding

The primary study was funded by WHO and partly by CDC. The funders did not play any role in the design, implementation, or scientific writing of this manuscript and did not influence the approval of this manuscript nor the choice of the journal or publication.

## Notes

### Competing Interest Statement

The authors have declared no competing interest.

